# CHARACTERIZATION OF GENETIC LOCI ASSOCIATED WITH ALLERGIC CONJUNCTIVITIS

**DOI:** 10.1101/2025.05.16.25327739

**Authors:** Fredrika Koskimäki, Oona Ahokas, Johanna Liinamaa, Kadri Reis, Anu Reigo, FinnGen, Estonian Biobank Research Team, Priit Palta, Johannes Kettunen, Minna K. Karjalainen, Ville Saarela

**Author notes:** Corresponding author: Fredrika Koskimäki, MD, Department of Ophthalmology and Medical Research Center, Oulu University Hospital; Research Unit of Clinical Medicine, University of Oulu, Oulu, Finland, Mail address: Kiviharjuntie 11, 90220 Oulu University Hospital, Oulu, Finland. These authors contributed equally to this work as last author (Minna K. Karjalainen, Ville Saarela).

## Abstract

**Importance:** Allergic conjunctivitis, a common presentation of allergy, co-occurs frequently with allergic rhinitis, atopic eczema and asthma. Despite its high prevalence, the genetic factors contributing to allergic conjunctivitis have not been characterized in detail.

**Objective:** We aimed to characterize genetic factors associated with allergic conjunctivitis both in relation to and independent of systemic atopic or allergic conditions through genome-wide association study (GWAS) meta-analysis.

**Design, setting and participants:** We performed a GWAS meta-analysis of allergic conjunctivitis utilizing data from FinnGen, the Estonian biobank and the UK biobank. In total 45 734 cases with allergic conjunctivitis and 1 084 159 controls were included. To get insights into allergic conjunctivitis-associated loci, we conducted a phenome-wide association study, performed pathway and enrichment analyses, and assessed genetic correlations with other phenotypes.

**Main outcomes and measures:** Genetic variants associated with allergic conjunctivitis.

**Results:** Genome-wide significant (*p* < 5 × 10^-8^) associations were identified for allergic conjunctivitis at 34 loci, many of which had not been reported to associate with allergic conjunctivitis before. Many of the associated loci included genes involved in immunology and allergy-related conditions, e.g., *ID2* (OR = 0.94, *p* = 2.5 × 10^-15^) and *TSLP* (OR = 1.06, *p* = 2.5 × 10^-14^). Several loci associated with allergic conjunctivitis were also associated with other allergic health conditions, such as asthma, allergic rhinitis and eczema. Three loci (*EIF2AK2*, *RANBP2*, *NFAT5*) had no previous association to allergy-related phenotypes. In pathway analysis, we detected that allergic conjunctivitis is associated with pathways related to neuroinflammation, immune responses and cytokine signaling, including those involved in T cell activation and differentiation. We detected an enrichment of genes associated with immune-related biological processes such as cytokine production and inflammatory response. Allergic conjunctivitis was genetically correlated with 27 phenotypes, including, e.g., doctor diagnosed hay fever or allergic rhinitis (*r_g_*=0.82).

**Conclusions and relevance:** We identified 34 allergic conjunctivitis associated loci, majority of which were involved in immunology and allergy-related conditions. Our findings underline the central role of inflammation-related genes in genetic predisposition to allergic conjunctivitis and advance the overall understanding of its genetic background.

## INTRODUCTION

Allergic conjunctivitis, characterized by an inflammatory response of the conjunctiva to an allergen, is a common presentation of allergy in both pediatric and adult population with an increasing global prevalence.^1^ It is most often induced by immunoglobulin E (IgE)-mediated mast cell degranulation, leading to the release of mediators such as histamine and cytokines.^2^ However, it may also result from a T-lymphocyte-mediated immune response or be part of a larger systemic atopic reaction.^3^ Typical symptoms include bilateral redness, itching, conjunctival chemosis and watery discharge.^4^ It is frequently associated with allergic rhinitis but it can be associated also with other conditions including dry eye disease, atopic dermatitis and asthma.^5–8^ Due to its high prevalence, it has a substantial influence on both quality of life and economic burden experienced by the patients.^9–11^

The heritability for allergic disease has been reported to be as high as 33-91% for allergic rhinitis and 71-84% for atopic dermatitis.^12^ While the genetics of various allergic conditions, such as allergic rhinitis^13^, have been studied extensively, the loci associated with allergic conjunctivitis have not been characterized in detail. Previously, a genome-wide association study (GWAS) for inflammatory and infectious upper respiratory diseases identified a locus associated with nasal polyposis, vasomotor and allergic rhinitis and chronic rhinosinusitis near *IL18RAP* which colocalized with allergic conjunctivitis.^14^ Another novel vasomotor and allergic rhinitis associated locus, *EMSY*, was also found to colocalize with allergic conjunctivitis.^14^

Despite its high prevalence, the genetic factors contributing to allergic conjunctivitis have not been investigated in detail. Here, we aimed to characterize the loci associated with allergic conjunctivitis both in relation to and independent of systemic atopic or allergic conditions.

## MATERIALS AND METHODS

### Study populations

This GWAS meta-analysis for allergic conjunctivitis included 1 129 893 participants of European descent only, with 45,734 cases with allergic conjunctivitis and 1 084 159 controls. Three large biobank-related studies were included: the FinnGen study^15^ with 29 791 allergic conjunctivitis cases and 470 557 controls, the Estonian Biobank (ESTBB)^16^ with 12 154 cases and 196 849 controls, and the UK Biobank (UKBB)^17^ with 3789 cases and 416 753 controls. The FinnGen study is a large-scale genomics initiative that has analyzed over 500 000 Finnish biobank samples and correlated genetic variation with health data to understand disease mechanisms and predispositions.^15^ The project is a collaboration between research organizations and biobanks within Finland and international industry partners. The present study included data from FinnGen Data Release R12. The Estonian Biobank is a population-based biobank with 212 955 participants in the current data freeze (2023v4).^16^ The UK Biobank is a large-scale open database including a half million individuals with paired genetic and phenotype information that has been enormously valuable in studies of genetic etiology for common diseases and traits.^17^ Detailed study descriptions are available in Supplemental Text.

### Definition of allergic conjunctivitis

In the FinnGen, allergic conjunctivitis cases were defined through hospital discharge registries using the ICD10 code H10.1, and through Kela registry, where the use of eye anti-allergy eye drops (ATC - S01GX) indicated allergic/atopic conjunctivitis. In both the ESTBB and in the UKBB, patients with ICD10 code H10.1 and ATC in S01GX were included as cases.

### Genome-wide association study and meta-analysis

GWAS was performed in each study, and the studies were combined in a meta-analysis. Mixed-model-based whole-genome regression with the REGENIE pipeline^18^ was used for GWAS in FinnGen, including sex, age, ten principal components (PC) and genotyping batches as covariates. REGENIE, v2.2.4^18^ with standard binary trait settings was used for GWAS in the ESTBB; variants with an INFO score > 0.4 and minor allele count > 2 were included, and age, age², sex and 10 PCs were included as covariates. In the UKBB, association analysis was performed using Scalable and Accurate Implementation of Generalized mixed model (SAIGE) with age, sex, age*sex, age²,age²*sex and 10 PCs as covariates for variants with an INFO score > 0.8 and minor allele count >20^19^. Meta-analysis was performed using an inverse variance weighted fixed-effect meta-analysis. Single nucleotide polymorphisms (SNPs) with *p*-values < 5 × 10^−8^ were considered significant, with the lead SNPs defined as the most significant SNP within each locus having a distance of at least 1 Mb. Manual curation and literature review was conducted to identify the most biologically plausible candidate genes at each locus using relevant databases including PubMed^20^, UniProt^21^ and Gene^22^. Genomic positions are for human genome build GRCh38.

### Comparison to previous associations

Novel loci for allergic conjunctivitis were defined as those more than 1 Mb away from previously reported associations.^14^ We compared the loci to previous allergic phenotypes in FinnGen, including allergic asthma, allergic rhinitis, asthma and allergy, childhood allergy (age <16), pollen allergy, allergic contact dermatitis, allergic urticaria, allergic contact dermatitis due to drugs in contact with skin, allergic purpura, asthma/COPD (KELA code 203), asthma (more control exclusions), asthma (only as main-diagnosis) (more control exclusions), suggestive for eosinophilic asthma, and atopic dermatitis. We also compared the loci to previous allergic associations in previous literature within 1Mb of lead SNPs, and in the NHGRI-EBI Catalog of human genome-wide association studies (GWAS catalog)^23^ through VannoPortal (http://www.mulinlab.org/vportal/index.html).

### Phenome-wide association study

A phenome-wide association study (PheWAS) was conducted to explore associations of allergic conjunctivitis-associated loci with across various phenotypes and traits. The PheWAS was performed using the GWAS catalog^23^ through VannoPortal (http://www.mulinlab.org/vportal/index.html); and by investigating associations across FinnGen^15^ R12 endpoints. Additionally, we assessed associations with gene expression levels using VannoPortal. Associations with *p*-value < 5 × 10^−8^ are reported.

### Pathway analyses

We performed pathway analyses to explore the association of allergic conjunctivitis with biological pathways utilizing the Functional Mapping and Annotation of Genome-Wide Association Studies (FUMA) GWAS platform.^24^ We performed MAGMA gene set analysis (SNP2GENE) and gene set enrichment analysis (GENETOFUNC) as implemented in FUMA. Details are shown in Supplementary Text.

### Genetic correlations

Linkage disequilibrium score regression (LDSC)^25^ was utilized to estimate genetic correlations, with significance set at p < 0.000168 after Bonferroni correction for 298 tests. We included 298 traits that were categorized into following groups: diseases, eye and ear related, cardiovascular, neurology, lung function, pain (skeletal), mood behavior, anthropometry and smoking. GWAS summary statistics from the MRC IEU OpenGWAS database were used.^26^

## RESULTS

### Genome-wide association study

In the GWAS meta-analysis, we identified 34 loci associated with allergic conjunctivitis (Figure 1, Table 1, Supplementary Table S1). Two of the loci (*IL18RAP*, *EMSY*) were previously described in relation to allergic conjunctivitis^14^, but the majority were novel.

**Figure 1.**
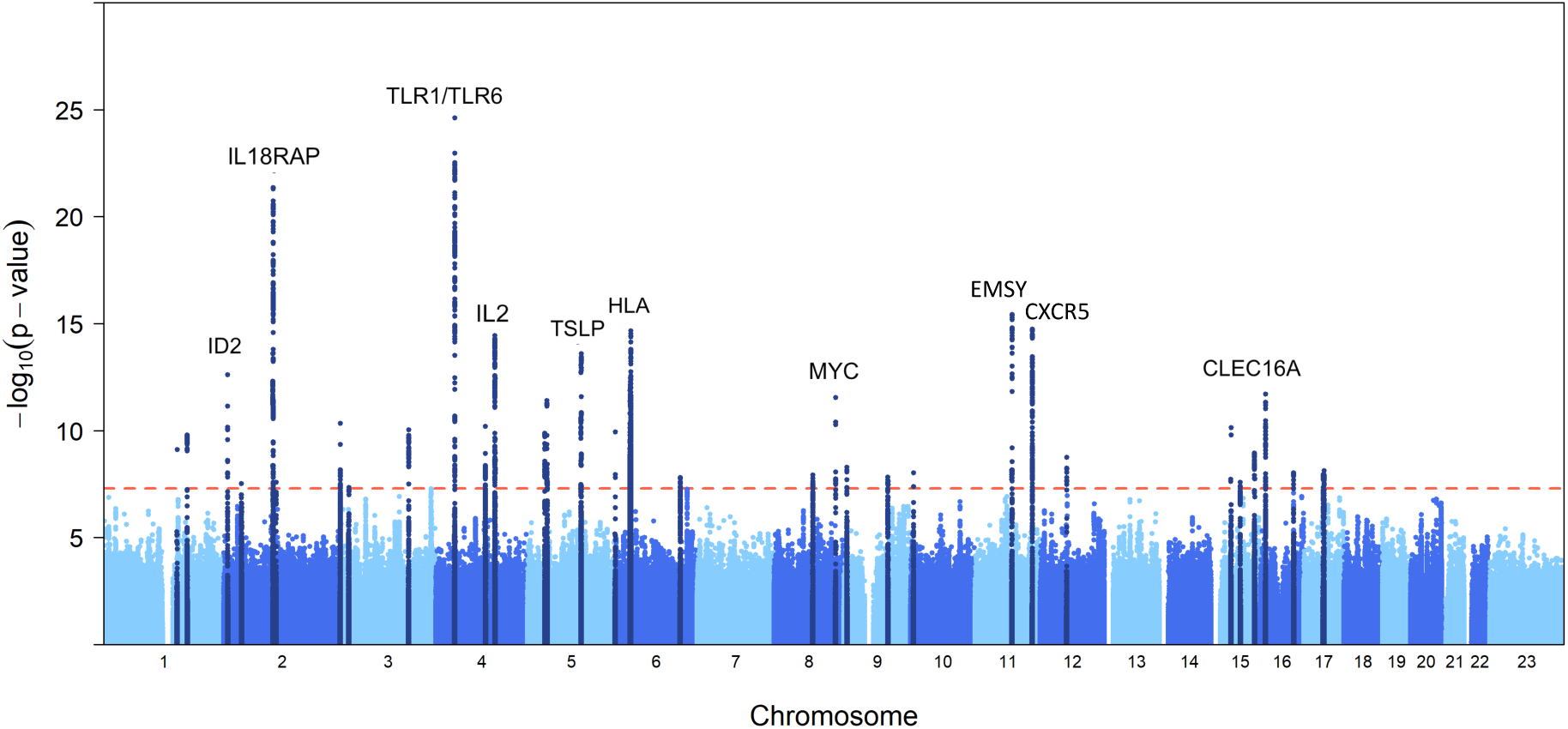
Manhattan plot showing the results of genome-wide association study of allergic conjunctivitis. Each dot represents a genetic variant. Chromosomal positions are shown on the X-axis, and Y-axis shows the association p-values on the -log10 scale. The genome-wide significance threshold is indicated with a red line. The candidate genes are indicated for the most significant association signals.

**Table 1.** Lead single nucleotide polymorphisms at loci associated with allergic conjunctivitis.

| Genomic position | rsID | Reference allele/<br>Effect allele | Effect allele frequency<br>FinnGen/ESTBB/UKBB | OR (95% CI) | p-value | Nearest/Candidate gene |
| --- | --- | --- | --- | --- | --- | --- |
| 1:152312600 | rs558269137 | CATG/C | 0.014/0.017/NA | 1.21 (1.14-1.28) | $7.76 \times 10^{-10}$ | *FLG* |
| 1:173182897 | rs6691738 | T/G | 0.300/0.277/0.285 | 1.05 (1.03-1.06) | $1.65 \times 10^{-10}$ | *TNFSF4* |
| 2:8302417 | rs891058 | G/A | 0.346/0.271/0.295 | 0.94 (0.93-0.96) | $2.53 \times 10^{-15}$ | *ID2* |
| 2:36960997 | rs7242096 | A/C | 0.302/0.247/0.255 | 1.04 (1.03-1.06) | $3.01 \times 10^{-8}$ | *EIF2AK2* |
| 2:102319699 | rs72823641 | T/A | 0.159/0.175/0.137 | 0.91 (0.90-0.93) | $4.34 \times 10^{-22}$ | *IL18RAP* |
| 2:108746109 | rs12614691 | G/A | 0.059/0.023/0.0085 | 0.91 (0.88-0.94) | $2.66 \times 10^{-8}$ | *RANBP2\** |
| 2:241759225 | rs34290285 | G/A | 0.216/0.296/0.256 | 0.95 (0.93-0.96) | $4.67 \times 10^{-11}$ | *PDCD1* |
| 3:17411433 | rs1446300 | C/A | 0.640/0.667/0.685 | 1.04 (1.03-1.06) | $4.63 \times 10^{-8}$ | *TBC1D5\** |
| 3:141374443 | rs7624084 | T/C | 0.446/0.396/0.443 | 1.05 (1.03-1.06) | $9.13 \times 10^{-11}$ | *ZBTB38* |
| 4:38797027 | rs5743618 | C/A | 0.153/0.212/0.226 | 0.91 (0.89-0.92) | $2.41 \times 10^{-25}$ | *TLR1/TLR6* |
| 4:102625698 | rs12645469 | G/A | 0.298/0.292/0.275 | 0.95 (0.94-0.97) | $6.4 \times 10^{-11}$ | *NFKB1* |
| 4:122307977 | rs7688384 | C/T | 0.311/0.334/0.316 | 0.94 (0.93-0.96) | $3.67 \times 10^{-15}$ | *IL2* |
| 5:35879054 | rs6881706 | G/T | 0.331/0.282/0.275 | 0.95 (0.94-0.97) | $1.31 \times 10^{-10}$ | *IL7R* |
| 5:40492553 | rs7714574 | C/T | 0.511/0.514/0.603 | 1.05 (1.03-1.06) | $4.01 \times 10^{-12}$ | *PTGER4* |
| 5:111092097 | rs79881291 | C/T | 0.311/0.281/0.361 | 1.06 (1.04-1.07) | $2.5 \times 10^{-14}$ | *TSLP* |
| 6:382897 | rs9378774 | A/G | 0.633/0.589/0.724 | 1.04 (1.03-1.06) | $1.16 \times 10^{-10}$ | *IRF4* |
| 6:31358354 | rs73390208 | C/T | 0.307/NA/0.267 | 0.94 (0.92-0.95) | $1.38 \times 10^{-12}$ | *HLA-B/HLA-C* |
| 6:32635544 | rs62404084 | C/T | 0.294/NA/0.193 | 0.93 (0.91-0.95) | $2.22 \times 10^{-15}$ | *HLA-DQA1/HLA-DQB1* |
| 6:135360566 | rs2614276 | T/C | 0.614/0.620/0.608 | 0.96 (0.95-0.97) | $1.6 \times 10^{-8}$ | *AHI1\** |
| 8:80384604 | rs13272105 | C/A | 0.655/NA/0.613 | 0.95 (0.94-0.97) | $1.22 \times 10^{-8}$ | *ZBTB10* |
| 8:127765520 | rs13279820 | T/C | 0.729/0.756/0.727 | 0.95 (0.93-0.96) | $2.86 \times 10^{-12}$ | *MYC* |
| 9:6091477 | rs1510077709 | T/TAAAC | 0.772/NA/0.760 | 1.06 (1.04-1.08) | $5.16 \times 10^{-9}$ | *IL33* |
| 9:91178108 | rs2478963 | T/C | 0.613/0.667/0.648 | 0.96 (0.95-0.97) | $1.54 \times 10^{-8}$ | *AUH\** |
| 10:6052734 | rs61839660 | C/T | 0.041/0.052/0.099 | 1.09 (1.06-1.13) | $9.81 \times 10^{-9}$ | *IL2RA* |
| 11:76582682 | rs7936312 | G/T | 0.414/0.444/0.477 | 1.06 (1.04-1.07) | $3.7 \times 10^{-16}$ | *EMSY* |
| 11:118809336 | rs2508569 | C/G | 0.685/0.675/0.658 | 1.06 (1.05-1.08) | $1.83 \times 10^{-15}$ | *CXCR5* |
| 12:55156742 | rs190653364 | G/A | 0.0064/0.0014/0.00095 | 1.33 (1.21-1.46) | $1.84 \times 10^{-9}$ | *TESPA1* |
| 15:41490486 | rs12440045 | A/C | 0.518/0.580/0.528 | 1.05 (1.03-1.06) | $7.35 \times 10^{-11}$ | *TYRO3* |
| 15:60776755 | rs11071557 | T/C | 0.154/0.189/0.130 | 0.95 (0.93-0.97) | $2.61 \times 10^{-8}$ | *RORA* |
| 15:90472334 | rs4031435 | G/C | 0.328/0.291/0.324 | 0.96 (0.94-0.97) | $1.16 \times 10^{-9}$ | *IQGAP1* |
| 16:11116558 | rs9923856 | T/C | 0.265/0.313/0.307 | 0.95 (0.93-0.96) | $2.00 \times 10^{-12}$ | *CLEC16A* |
| 16:69702647 | rs10714023 | GC/G | 0.257/0.249/0.266 | 0.96 (0.94-0.97) | $9.51 \times 10^{-9}$ | *NFAT5* |
| 17:40619224 | rs9911533 | C/T | 0.606/0.593/0.638 | 1.04 (1.03-1.06) | $1.18 \times 10^{-8}$ | *CCR7* |
| 17:42445537 | rs36005199 | A/G | 0.286/0.238/0.273 | 0.96 (0.94-0.97) | $7.8 \times 10^{-9}$ | *STAT3* |

rsID, reference SNP cluster ID; OR, odds ratio; 95% CI, 95% confidence interval; ESTBB, Estonian BioBank; UKBB, UK BioBank; NA, not applicable, SNP not present in the data. Most significant single nucleotide polymorphism within each associated region is shown. Genomic position is presented as chromosome:position. When no biologically plausible gene was identified, the nearest gene is shown and marked with \*.

We identified biologically plausible genes in most of the associated loci (Table 1). The most significant association was near the toll-like receptor 1 (*TLR1*) and toll-like receptor 6 (*TLR6*) genes (rs5743618, *p* = 2.4 ×10^-25^). At this locus, the strongest association was observed in FinnGen (OR 0.90, *p* = 9.5× 10^-19^). In the Estonian Biobank, this association was replicated with a similar effect as in FinnGen (OR = 0.90 and *p* = 4.2×10^-9^), while the association in the UK Biobank was not significant (OR = 0.97 and *p* = 0.23) (Supplementary Table S1, Supplementary Figure S1). Toll-like receptors recognize specific pathogen associated molecular patterns that are present on the surface of pathogens, and are fundamental in innate immunity activation.^27^ This region has been previously associated with self-reported allergy, allergic sensitization and the risk of having asthma with hay fever.^28–30^

Overall, the majority of the associated loci had roles in immunology and allergy-related conditions such as allergic rhinitis, eczema and asthma. Regional association plots for exemplary loci are shown in Supplementary Figure S2. For example, we detected associations at 5 loci containing genes that encode interleukin receptors or interleukins including the *IL18RAP*, *IL2*, *IL7R*, *IL33* and *IL2RA*/*IL15RA* loci. Associations were also identified at close proximity of chemokine receptor coding genes (*CCR7*, *CXCR5*). One of the association signals was near the gene encoding inhibitor of DNA binding 2 (*ID2*) (lead SNP rs891058, *p* = 2.5 × 10^-15^). In murine models, ID2 reduces class switch recombination to immunoglobulin E (IgE) to prevent serious complications such as allergic diseases or anaphylaxis.^31^ It has been suggested that genetic variants within the *ID2* gene may be present in syndromes characterized by elevated IgE levels, such as allergic conditions.^31^ Another association near the thymic stromal lymphopoietin (*TSLP*) gene was identified (lead SNP rs79881291, *p* = 2.5 × 10^-14^). Epithelial cytokine TSLP is recognized for its capacity to activate T helper type 2 (Th2) immune responses against extracellular antigens and it plays a crucial role in the development of allergic inflammation.^32^ In previous studies, the expression of TSLP has been elevated in conjunctiva of patients with different types of allergic conjunctivitis compared to controls, demonstrating that it may contribute to development and progression of allergic conjunctivitis.^33^

At most loci, the strongest associations were observed in the FinnGen cohort, due to its large size, with similar effects replicated in the ESTBB and the UKBB (Supplementary Figure S1). For example, SNP rs79881201 near *TSLP* shows consistent effect across the FinnGen (OR 1.06, *p* = 1.6× 10^-10^), the ESTBB (OR 1.04, *p* = 3.3× 10^-3^) and the UKBB (OR 1.08, *p* = 1.6× 10^-3^). In contrast, rs190653364 near *TESPA1* demonstrates variability, with similar effects observed in the FinnGen and ESTBB cohorts (OR = 1.31 and OR = 1.61, respectively) but no association in the UKBB (OR = 0.90, *p* = 0.82).

### Phenome-wide association study

We performed a phenome-wide association study of the loci associated with allergic conjunctivitis to investigate the associations across phenotypes including diseases and traits, and gene expression levels (Supplementary Table S2, S3 and S4). We found that several SNPs displayed associations with different immune-related phenotypes such as asthma, allergic rhinitis and eczema. However, the lead SNP rs7624084 near the *ZBTB38* gene associated with eye-related conditions including spherical equivalent and myopia (age at diagnosis), rather than with immune-related disease.

We studied the associations of allergic conjunctivitis loci against all predetermined FinnGen endpoints (Supplementary Table S4). Many of the loci demonstrated associations with immune-related phenotypes including association with asthma, allergic asthma or childhood asthma in 16 loci, and with atopic dermatitis in 14 loci. We also detected associations with eye-related conditions including anterior uveitis, and diabetic retinopathy and maculopathy.

We also studied associations of the lead SNPs with gene expression levels (Supplementary Table S3). We detected that twelve of the lead SNPs were associated with gene expression levels of the candidate genes, such as the lead SNP rs79881201 near the *TSLP* gene that was associated with the expression levels of the *TSLP* in various tissues including testis, tibial artery, subcutaneous adipose tissue and esophagus mucosa.

### Comparison to previous allergic conjunctivitis associations and allergy-related phenotypes

There were no loci associated with allergic conjunctivitis in GWAS Catalog. However, two genetic loci were previously described in relation to allergic conjunctivitis (Figure 1, Table 1, Supplementary Table S1).^14^

These included the *IL18RAP* (interleukin 18 receptor accessory protein; rs72823641, *p* = 4.3 × 10^-22^) and *EMSY* (EMSY transcriptional repressor, BRCA2 interacting; rs7936312, *p* = 3.7 × 10^-16^) loci.

To investigate whether there are susceptibility loci that have not been previously reported to be associated with allergic disease, we explored associations of the allergic conjunctivitis associated loci with selected allergy-related phenotypes within 1 Mb of the lead SNPs. Most of the loci had allergy-related associations in literature (Supplementary Table S2, S4 and S7). Interestingly, we detected that three of the loci (*EIF2AK2*, *RANBP2* and *NFAT5*) had no previous associations related to allergic diseases. At least two of the loci, *EIF2AK2* and *NFAT5*, have genes with roles in immunity. Eukaryotic translation initiation factor 2 alpha kinase 2 gene, *EIF2AK2*, has a central role in the antiviral immune response and it has been implicated in acute exacerbations in asthmatics.^34^ *EIF2AK2* has been reported to associate with high light scatter reticulocyte percentage of red cells.^35^ Nuclear factor of activated T cells 5 gene, *NFAT5*, is involved in the regulation of immune response and it may have a role in inflammation in dry eye disease.^36^ The nearest gene in the third locus, *RANBP2*, encoding RAN binding protein 2, is involved in DNA replication, and in the regulation of nucleocytoplasmic transportation.^37^ Other genes at this locus include *LIMS1*, *CCDC138* and *EDAR* (Supplementary Figure S2).

### Pathway analyses

In pathway analyses, we detected significant associations with pathways related to neuroinflammation, immune responses and cytokine signaling (Table 2). Specific pathways included those involved in neuroinflammation and glutamatergic signaling, NAD+ nucleosidase activity, cytokine-cytokine receptor interactions, and T cell activation and differentiation. Pathways related to COVID-19 and regulatory T cell function were also identified. In the gene enrichment analysis, we detected an enrichment of genes associated with immune-related biological processes such as cytokine production and inflammatory response (Supplementary Table S6).

**Table 2.** Magma gene set analysis.

| Gene set | Number of genes in the gene set | Bonferroni corrected p-value |
| --- | --- | --- |
| WP NEUROINFLAMMATION AND GLUTAMATERGIC SIGNALING | 138 | $1.54 \times 10^{-4}$ |
| GOMF NADPLUS NUCLEOSIDASE ACTIVITY | 16 | $8.18 \times 10^{-4}$ |
| KEGG CYTOKINE CYTOKINE RECEPTOR INTERACTION | 249 | $1.67 \times 10^{-3}$ |
| WP FOXP3 IN COVID19 | 15 | 0.0112 |
| GOBP CD4 POSITIVE ALPHA BETA T CELL ACTIVATION | 112 | 0.0150 |
| GOBP REGULATION OF T HELPER 1 TYPE IMMUNE RESPONSE | 30 | 0.0244 |
| GOBP T CELL DIFFERENTIATION INVOLVED IN IMMUNE RESPONSE | 78 | 0.0352 |
| WP TH17 CELL DIFFERENTIATION PATHWAY | 68 | 0.0388 |
| GPBD CD4 POSITIVE ALPHA BET T CELL DIFFERENTIATION | 87 | 0.0410 |

### Genetic correlations

We performed a genetic correlation analysis of allergic conjunctivitis with 298 traits (Supplementary Table S5) and altogether 27 genetic correlations were found significant (*p* < 0.000168, Figure 2). In the context of allergic diseases, a high genetic correlation between allergic conjunctivitis and doctor diagnosed hay fever or allergic rhinitis was detected (*r_g_*=0.82, *p* = 2.8 × 10^-80^) as well as a moderate genetic correlation with eczema (*r_g_*=0.50, *p* = 6.7 × 10^-9^). A moderate genetic correlation was identified between allergic conjunctivitis and wheeze or whistling in the chest last year (*r_g_*=0.31, *p* = 8.0 × 10^-19^), chest pain or discomfort (*r_g_*=0.23, *p* = 5.1 × 10^-10^) and lung function (FEV1/FVC) (*r_g_*=-0.11, *p* = 1.9 × 10^-5^) and other eye problems (*r_g_*=0.24, *p* = 8.1 × 10^-6^). In addition, various nominal genetic correlations were identified (Supplementary Table S5).

**Figure 2.**
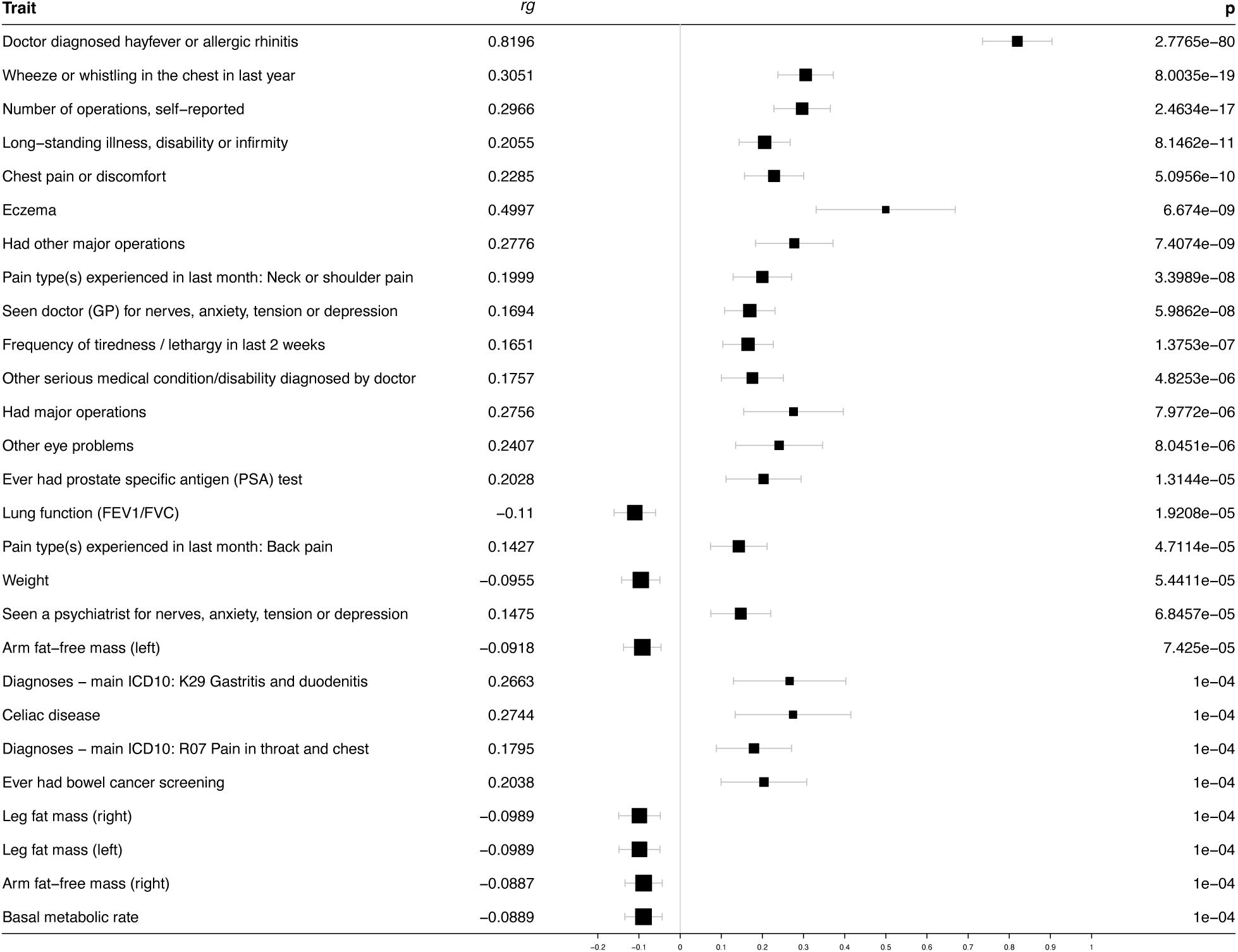
Forest plot displaying genetic correlations (*r_g_*) of allergic conjunctivitis with other diseases and traits. Genetic correlation analysis identified 27 traits with genetic correlations with allergic conjunctivitis (*p* < 0.000168).

We also investigated the proportion of allergic conjunctivitis cases having other allergy-related health conditions in FinnGen. Of all allergic conjunctivitis cases, 22% also had asthma (as main diagnosis) (Jaccard index 9.4%), 18% had atopic dermatitis (Jaccard 9.4%), 15% had allergic rhinitis (Jaccard 10.8%), 11% had asthma and allergy (Jaccard 9.6%), 11% had pollen allergy (Jaccard 9.1%) and 11% had allergic asthma (Jaccard 8.0%). In contrast, conditions such as allergic contact dermatitis, eosinophilic asthma, drug-related allergic contact dermatitis, allergic purpura, and food allergy showed minimal overlap (<3%).

## DISCUSSION

In this genome-wide association study, we characterized 34 allergic conjunctivitis associated loci in detail for the first time. Majority of the loci associated with allergic conjunctivitis, including e.g. *TLR1*/*TLR6*, *ID2* and *TSLP*, had genes involved in immunology and allergic conditions such as allergic rhinitis, eczema and asthma. Interestingly, three of the identified loci had no previous allergy-related associations. We further detected that many of the loci associated with allergic conjunctivitis also showed associations with other immune-related conditions, such as asthma, allergic rhinitis and eczema. In addition, we detected genetic correlations between allergic conjunctivitis and diagnosed hay fever or allergic rhinitis, eczema, wheeze or whistling in the chest last year, chest pain or discomfort, lung function and other eye problems.

We detected that three of the identified genetic loci, *EIF2AK2*, *RANBP2* and *NFAT5*, have no previous allergy-related associations. These loci emerge as intriguing candidates with broader implications in allergy-related processes. *EIF2AK2* is a key regulator of the antiviral immune response, with a well-established role in host defense against both DNA and RNA viruses.^34^ *EIF2AK2* has also been linked to acute asthma exacerbations.^34^ This dual role highlights its significance in both immune protection and inflammatory disease processes. *RANBP2* polymorphisms are known to influence susceptibility to pediatric gliomas.^37^ The potential roles of *RANBP2* and other genes at this locus in allergic conjunctivitis remain unclear and present an interesting opportunity for future research. One of the loci, *NFAT5*, has been previously linked to dry eye disease.^36^ Both allergic conjunctivitis and dry eye disease involve inflammation that affects the ocular surface, and they often co-occur^5^, highlighting the potential role of *NFAT5* in ocular surface disease. *NFAT5* also associates with height, which is of interest since growth impairment has been reported in children with allergies.^38^ However, underlying mechanisms are not well understood, and it has been suggested that growth impairment does not persist to adulthood.^38^ Although these are seemingly distinct conditions, further studies could illuminate possible shared biological mechanisms that bridge allergic conjunctivitis with other traits. The specific roles of these three loci in allergic conjunctivitis remain to be studied further. In our phenome-wide association study, we detected that many of the allergic conjunctivitis associated loci also showed associations with other allergic conditions. Interestingly, the lead SNP rs7624084 near the *ZBTB38* gene was associated with eye-related conditions, including spherical equivalent and myopia (age of diagnosis), rather than with immune-related diseases. While the alternative allele C predisposed to allergic conjunctivitis, the allele T was associated with an increased risk of myopia at a younger age. This suggests a possible connection between allergic conjunctivitis and refractive error. In contrast, previous research has shown that children with allergic conjunctivitis are at a higher risk of developing myopia and moreover, animal studies indicate that allergic inflammation can promote myopia.^39^ Future studies could provide further insights into the underlying shared biology between allergic conjunctivitis and refractive errors.

Majority of the loci associated with allergic conjunctivitis have genes involved in immunity. This is not surprising, as for example allergic conjunctivitis and allergic rhinitis, though separate conditions, frequently co-occur. The degree of overlap varies, with allergic conjunctivitis manifesting in 52-65% of patients with allergic rhinitis.^40,41^ However, either condition can occur independently of the other.^42^ In contrast, less than twenty percent of allergic conjunctivitis cases were diagnosed with allergic rhinitis in FinnGen, indicating that individuals diagnosed with either condition partly represent distinct cases. The proportion of shared cases was even smaller for many other allergy-related conditions, such as pollen allergy and food allergy. Furthermore, as we identified allergic conjunctivitis associated loci that have not been reported to be associated with allergic disease before, the genetic factors behind the different allergy-related phenotypes appear to be at least partly nonoverlapping.

The large number of participants, with a total of 45 734 cases and 1 084 159 controls, is a significant strength of this study. This large size, and the inclusion of three separate cohorts, enables a more comprehensive identification of genetic factors associated with allergic conjunctivitis. From a clinical perspective, the diagnosis of allergic conjunctivitis tends to be generally reliable, enhancing the uniformity between cohorts. Despite these strengths, our study has certain limitations that need to be considered while interpreting our findings. The reliance on European cohorts restricts the generalizability of the findings, highlighting the need for future studies to include participants from diverse backgrounds. Nevertheless, this study provides significant novel insights into the genetic and biological mechanisms associated with allergic conjunctivitis.

## CONCLUSIONS

In this genome-wide association study, we identified 34 loci associated with allergic conjunctivitis. Majority of the loci demonstrated associations with immune-related phenotypes. Moreover, we detected genetic correlations between allergic conjunctivitis and allergy-related conditions, linking allergic conjunctivitis with broader atopic and immune-related pathways. Importantly, we also identified loci that had not been previously associated with allergic disease. Our findings advance our understanding of the genetic background of allergic conjunctivitis and the genetic connections between allergic conjunctivitis and atopic conditions, providing novel guidelines for future research.

## Data Availability

Summary statistics will be made publicly available upon publication.

## ACKNOWLEDGEMENTS

We want to acknowledge the participants and investigators of FinnGen study. The FinnGen project is funded by two grants from Business Finland (HUS 4685/31/2016 and UH 4386/31/2016) and the following industry partners: AbbVie Inc., AstraZeneca UK Ltd, Biogen MA Inc., Bristol Myers Squibb (and Celgene Corporation & Celgene International II Sàrl), Genentech Inc., Merck Sharp & Dohme LCC, Pfizer Inc., GlaxoSmithKline Intellectual Property Development Ltd., Sanofi US Services Inc., Maze Therapeutics Inc., Janssen Biotech Inc, Novartis Pharma AG, and Boehringer Ingelheim International GmbH. Following biobanks are acknowledged for delivering biobank samples to FinnGen: Auria Biobank (www.auria.fi/biopankki), THL Biobank (www.thl.fi/biobank), Helsinki Biobank (www.helsinginbiopankki.fi), Biobank Borealis of Northern Finland (https://www.ppshp.fi/Tutkimus-ja-opetus/Biopankki/Pages/Biobank-Borealis-briefly-in-English.aspx), Finnish Clinical Biobank Tampere (www.tays.fi/en-US/Research_and_development/Finnish_Clinical_Biobank_Tampere), Biobank of Eastern Finland (www.ita-suomenbiopankki.fi/en), Central Finland Biobank (www.ksshp.fi/fi-FI/Potilaalle/Biopankki), Finnish Red Cross Blood Service Biobank (www.veripalvelu.fi/verenluovutus/biopankkitoiminta), Terveystalo Biobank (www.terveystalo.com/fi/Yritystietoa/Terveystalo-Biopankki/Biopankki/) and Arctic Biobank (https://www.oulu.fi/en/university/faculties-and-units/faculty-medicine/northern-finland-birth-cohorts-and-arctic-biobank). All Finnish Biobanks are members of BBMRI.fi infrastructure (www.bbmri.fi). Finnish Biobank Cooperative -FINBB (https://finbb.fi/) is the coordinator of BBMRI-ERIC operations in Finland. The Finnish biobank data can be accessed through the Fingenious® services (https://site.fingenious.fi/en/) managed by FINBB. We want to acknowledge the participants of the Estonian Biobank for their contributions. The activities of the Estonian Biobank are regulated by the Human Genes Research Act, which was adopted in year 2000 specifically for the operations of the Estonian Biobank. Individual level data analysis in the Estonian Biobank was carried out under ethical approval “1.1-12/624” from the Estonian Committee on Bioethics and Human Research (Estonian Ministry of Social Affairs), using data according to release application “3-10/GI/1915” from the Estonian Biobank. The Estonian Genome Center GWAS analyses were performed in the High Performance Computing Center, University of Tartu. The work of the Estonian Genome Center, University of Tartu was funded by the Estonian Research Council Grant PRG1291. The authors also acknowledge CSC-IT Center for Science, Finland, for computational resources. This study has received funding from Oulu University Hospital VTR funding, the Silmäsäätiö Foundation, Suomen Lääketieteen Säätiö Foundation and the Mary and Georg C. Ehrnrooth Foundation. The funding organizations had no role in the design or conduct of this study. The authors have no conflict of interest.

