## Supplementary Text for "CHARACTERIZATION OF GENETIC LOCI ASSOCIATED WITH ALLERGIC CONJUNCTIVITIS"

Corresponding author:

Fredrika Koskimäki, MD

**SUPPLEMENTARY MATERIAL**

**Table of contents**

Supplementary Note: FinnGen DF12 ethics statement.....................................................................................2

Supplementary text: Detailed descriptions of study populations…………………………………………......3

Supplementary text: FUMA input parameters……..…………………………………………………………4

Figure S1. Forest plot displaying the effect estimates of lead single nucleotide polymorphisms……..……..5

Figure S2. Regional association plots of selected allergic conjunctivitis associated loci……………………9

Supplementary text: ESTBB consortium authors ………………………………………...…………………13

Supplemental material references ………………………………………………………………………...…14

**SUPPLEMENTARY NOTE: FinnGen DF12 ethics statement.**

Study subjects in FinnGen provided informed consent for biobank research, based on the Finnish Biobank Act. Alternatively, separate research cohorts, collected prior the Finnish Biobank Act came into effect (in September 2013) and start of FinnGen (August 2017), were collected based on study-specific consents and later transferred to the Finnish biobanks after approval by Fimea (Finnish Medicines Agency), the National Supervisory Authority for Welfare and Health. Recruitment protocols followed the biobank protocols approved by Fimea. The Coordinating Ethics Committee of the Hospital District of Helsinki and Uusimaa (HUS) statement number for the FinnGen study is Nr HUS/990/2017.

The FinnGen study is approved by Finnish Institute for Health and Welfare (permit numbers: THL/2031/6.02.00/2017, THL/1101/5.05.00/2017, THL/341/6.02.00/2018, THL/2222/6.02.00/2018, THL/283/6.02.00/2019, THL/1721/5.05.00/2019 and THL/1524/5.05.00/2020), Digital and population data service agency (permit numbers: VRK43431/2017-3, VRK/6909/2018-3, VRK/4415/2019-3), the Social Insurance Institution (permit numbers: KELA 58/522/2017, KELA 131/522/2018, KELA 70/522/2019, KELA 98/522/2019, KELA 134/522/2019, KELA 138/522/2019, KELA 2/522/2020, KELA 16/522/2020), Findata permit numbers THL/2364/14.02/2020, THL/4055/14.06.00/2020, THL/3433/14.06.00/2020, THL/4432/14.06/2020, THL/5189/14.06/2020, THL/5894/14.06.00/2020, THL/6619/14.06.00/2020, THL/209/14.06.00/2021, THL/688/14.06.00/2021, THL/1284/14.06.00/2021, THL/1965/14.06.00/2021, THL/5546/14.02.00/2020, THL/2658/14.06.00/2021, THL/4235/14.06.00/2021, Statistics Finland (permit numbers: TK-53-1041-17 and TK/143/07.03.00/2020 (earlier TK-53-90-20) TK/1735/07.03.00/2021, TK/3112/07.03.00/2021) and Finnish Registry for Kidney Diseases permission/extract from the meeting minutes on 4th July 2019.

The Biobank Access Decisions for FinnGen samples and data utilized in FinnGen Data Freeze 11 include: THL Biobank BB2017_55, BB2017_111, BB2018_19, BB_2018_34, BB_2018_67, BB2018_71, BB2019_7, BB2019_8, BB2019_26, BB2020_1, BB2021_65, Finnish Red Cross Blood Service Biobank 7.12.2017, Helsinki Biobank HUS/359/2017, HUS/248/2020, HUS/430/2021 §28, §29, HUS/150/2022 §12, §13, §14, §15, §16, §17, §18, §23, §58, §59, HUS/128/2023 §18, Auria Biobank AB17-5154 and amendment #1 (August 17 2020) and amendments BB_2021-0140, BB_2021-0156 (August 26 2021, Feb 2 2022), BB_2021-0169, BB_2021-0179, BB_2021-0161, AB20-5926 and amendment #1 (April 23 2020) and it´s modifications (Sep 22 2021), BB_2022-0262, BB_2022-0256, Biobank Borealis of Northern Finland_2017_1013, 2021_5010, 2021_5010 Amendment, 2021_5018, 2021_5018 Amendment, 2021_5015, 2021_5015 Amendment, 2021_5015 Amendment_2, 2021_5023, 2021_5023 Amendment, 2021_5023 Amendment_2, 2021_5017, 2021_5017 Amendment, 2022_6001, 2022_6001 Amendment, 2022_6006 Amendment, 2022_6006 Amendment, 2022_6006 Amendment_2, BB22-0067, 2022_0262, 2022_0262 Amendment, Biobank of Eastern Finland 1186/2018 and amendment 22§/2020, 53§/2021, 13§/2022, 14§/2022, 15§/2022, 27§/2022, 28§/2022, 29§/2022, 33§/2022, 35§/2022, 36§/2022, 37§/2022, 39§/2022, 7§/2023, 32§/2023, 33§/2023, 34§/2023, 35§/2023, 36§/2023, 37§/2023, 38§/2023, 39§/2023, 40§/2023, 41§/2023, Finnish Clinical Biobank Tampere MH0004 and amendments (21.02.2020 & 06.10.2020), BB2021-0140 8§/2021, 9§/2021, §9/2022, §10/2022, §12/2022, 13§/2022, §20/2022, §21/2022, §22/2022, §23/2022, 28§/2022, 29§/2022, 30§/2022, 31§/2022, 32§/2022, 38§/2022, 40§/2022, 42§/2022, 1§/2023, Central Finland Biobank 1-2017, BB_2021-0161, BB_2021-0169, BB_2021-0179, BB_2021-0170, BB_2022-0256, BB_2022-0262, BB22-0067, Decision allowing to continue data processing until 31st Aug 2024 for projects: BB_2021-0179, BB22-0067,BB_2022-0262, BB_2021-0170, BB_2021-0164, BB_2021-0161, and BB_2021-0169, and Terveystalo Biobank STB 2018001 and amendment 25th Aug 2020, Finnish Hematological Registry and Clinical Biobank decision 18th June 2021, Arctic biobank P0844: ARC_2021_1001.

**SUPPLEMENTARY TEXT: Detailed descriptions of study populations**

The FinnGen

The FinnGen study is a large-scale genomics initiative that has analyzed over 500,000 Finnish biobank samples and correlated genetic variation with health data to understand disease mechanisms and predispositions. The project is a collaboration between research organizations and biobanks within Finland and international industry partners.^1^ The present study included data from FinnGen Data Release R12.

The Estonian Biobank

The Estonian Biobank is a population-based biobank with 212,955 participants in the current data freeze (2023v4). All biobank participants have signed a broad informed consent form and information on ICD-10 codes is obtained via regular linking with the national Health Insurance Fund and other relevant databases, with majority of the electronic health records having been collected since 2004.^2^

All EstBB participants have been genotyped at the Core Genotyping Lab of the Institute of Genomics, University of Tartu, using Illumina Global Screening Array v1.0 and v2.0. Samples were genotyped, and PLINK format files were created using Illumina GenomeStudio v2.0.4. Individuals were excluded from the analysis if their call rate was <95% or if the sex defined based on heterozygosity of the X chromosome did not match the sex in phenotype data. Before imputation, variants were filtered by call rate <95%, Hardy–Weinberg equilibrium (HWE) P < 1 × 10−4 (autosomal variants only) and minor allele frequency <1%. Variant positions were updated to b37, and all variants were changed to be from TOP strand using GSAMD-24v1-0_20011747_A1-b37.strand.RefAlt.zip files from https://www.well.ox.ac.uk/~wrayner/strand/ webpage. Prephasing was done using Eagle v2.3 software38 (number of conditioning haplotypes Eagle2 uses when phasing each sample was set to:–Kpbwt=20000) and imputation was done using Beagle v.28Sep18.79339 with effective population size ne = 20,000. Population-specific imputation reference of 2,297 WGS samples was used.^3^

The activities of the EstBB are regulated by the Human Genes Research Act, which was adopted in 2000 specifically for the operations of EstBB. Individual level data analysis in EstBB was carried out under ethical approval nr 1.1-12/1020 from the Estonian Committee on Bioethics and Human Research (Estonian Ministry of Social Affairs), using data according to release application 3-10/GI/1915 from the Estonian Biobank.

The UK Biobank

The UK Biobank is a large-scale open database including a half million individuals with paired genetic and phenotype information that has been enormously valuable in studies of genetic etiology for common diseases and traits. The PanUKBB (<https://pan.ukbb.broadinstitute.org/>) project performed a pan-ancestry genetic analysis for 500 000 UK Biobank samples, with 40 to 69 years old adults as participants. The phenotypes were identified with registry data and questionnaires. The project is aiming to follow up participants for at least 30 years.^4^

**SUPPLEMENTARY TEXT: FUMA input parameters**

Lead SNPs were lifted to hg19 using liftOver tool (https://genome.ucsc.edu/cgi-bin/hgLiftOver). The GWAS summary statistics using rsID as identifier and predefined lead SNPs were imported to SNP2GENE platform. The maximum *p*-value <5 × 10^-8^ of lead SNPs was applied. Cut-off *p*-value <0.05 was used. The threshold to define independent significant SNPs was r^2^ ≥ 0.6 and second threshold to define lead SNPs was r^2^ ≥ 0.1. 1000G Phase3 EUR was used as the reference panel population. Maximum distance between LD blocks to merge into a single locus was 250 kb. Positional mapping was performed using functional consequences of SNPs on genes to map (exonic, splicing, intronic, 3UTR, 5UTR) with minimum CADD score of 12.37 and maximum RegulomeDB score 3b. HLA region was excluded from annotations in eQTL mapping. Tissue types GTExv8 blood, GTExv8 blood vessels and GTExv8 skin were included in the eQTL mapping. MAGMA gene expression analysis was performed with 10 kb gene window and GTExv8: 54 tissue types and GTExv8: 30 general tissue types. Mapped genes were uploaded to GENE2FUNC for a gene set enrichment analysis.

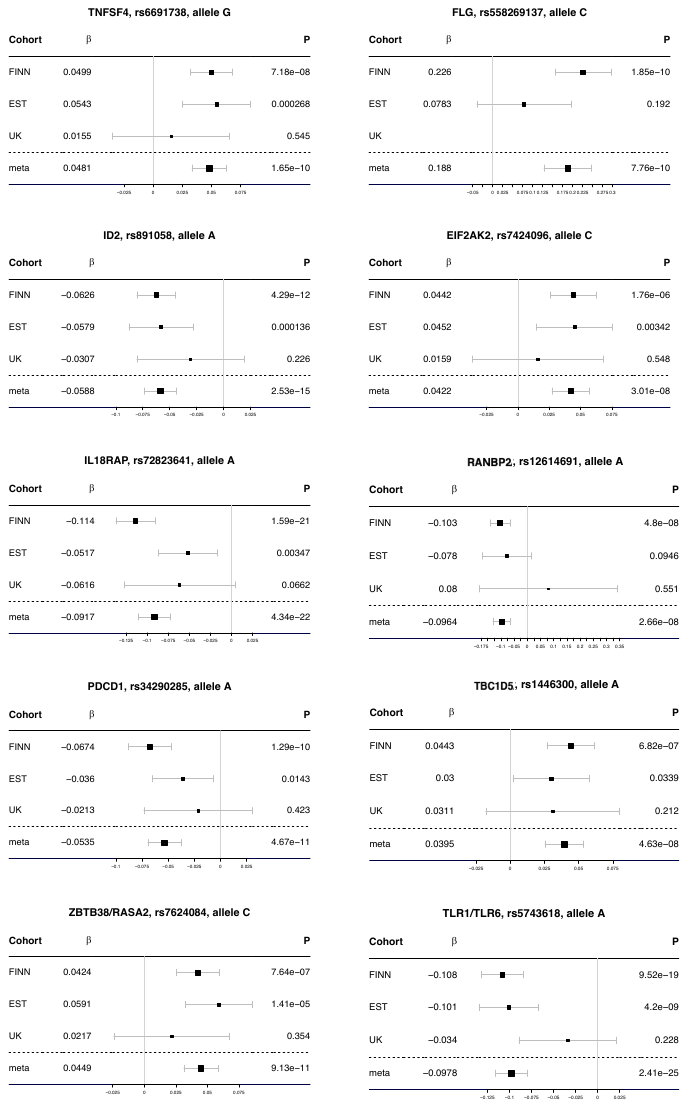

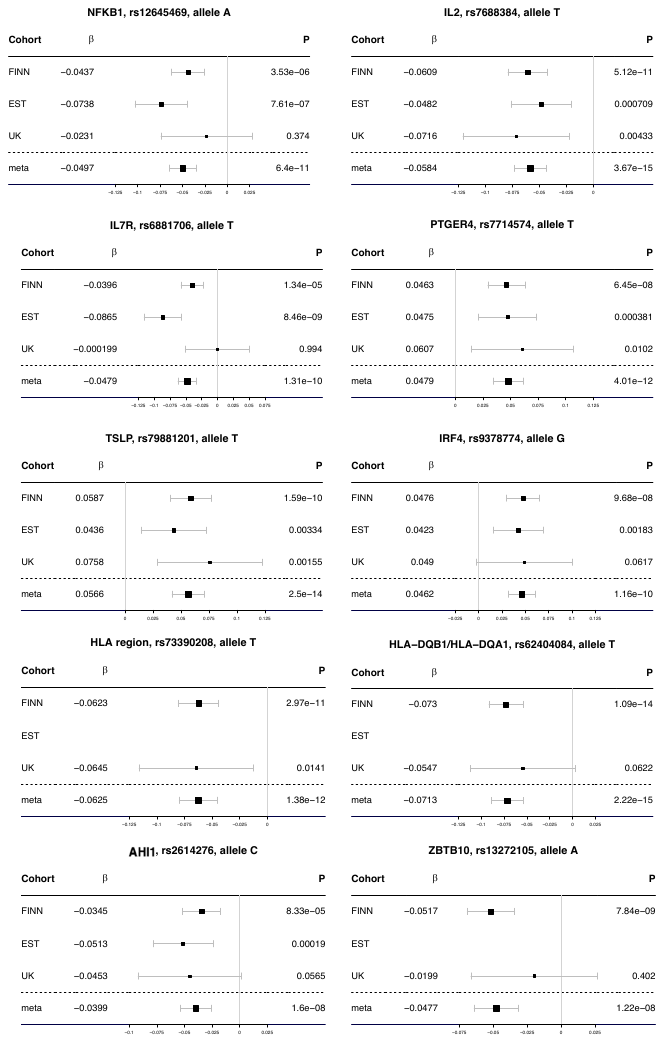

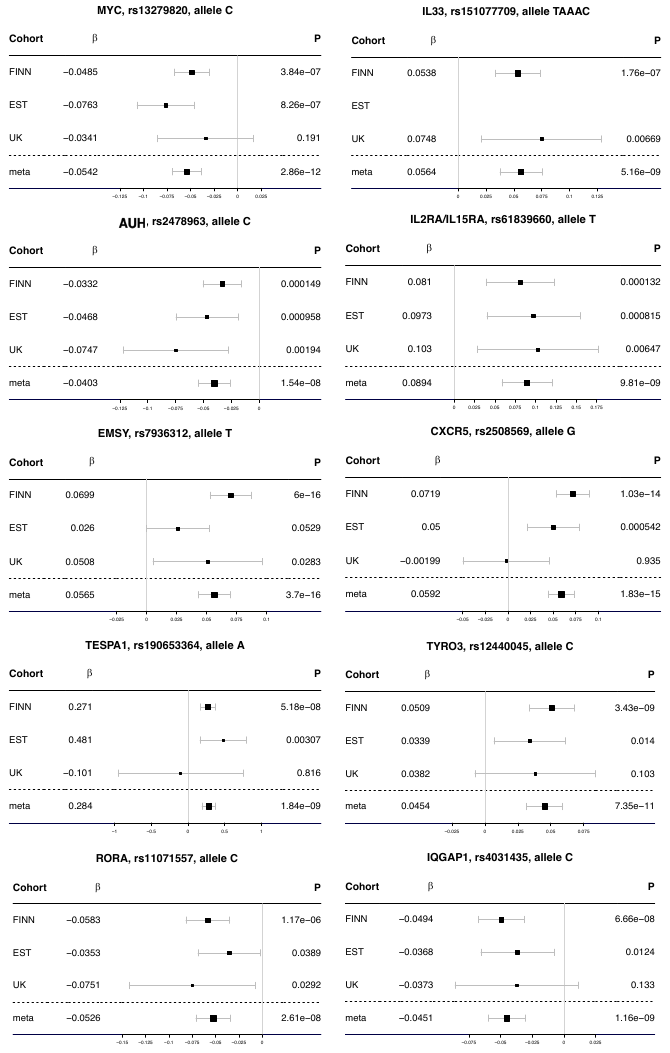

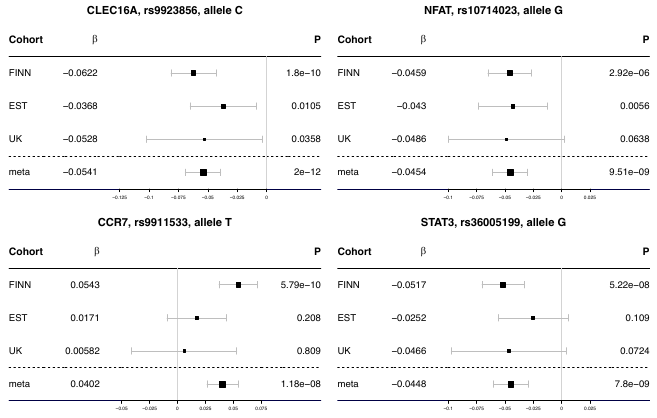

**Figure S1.** Forest plot displaying the effect estimates of lead single nucleotide polymorphisms at the loci associated with allergic conjunctivitis in each study population and meta-analysis. Whiskers are displaying the 95 % confidence interval. Finn, FinnGen; Est, Estonian Biobank; UK, UK Biobank.

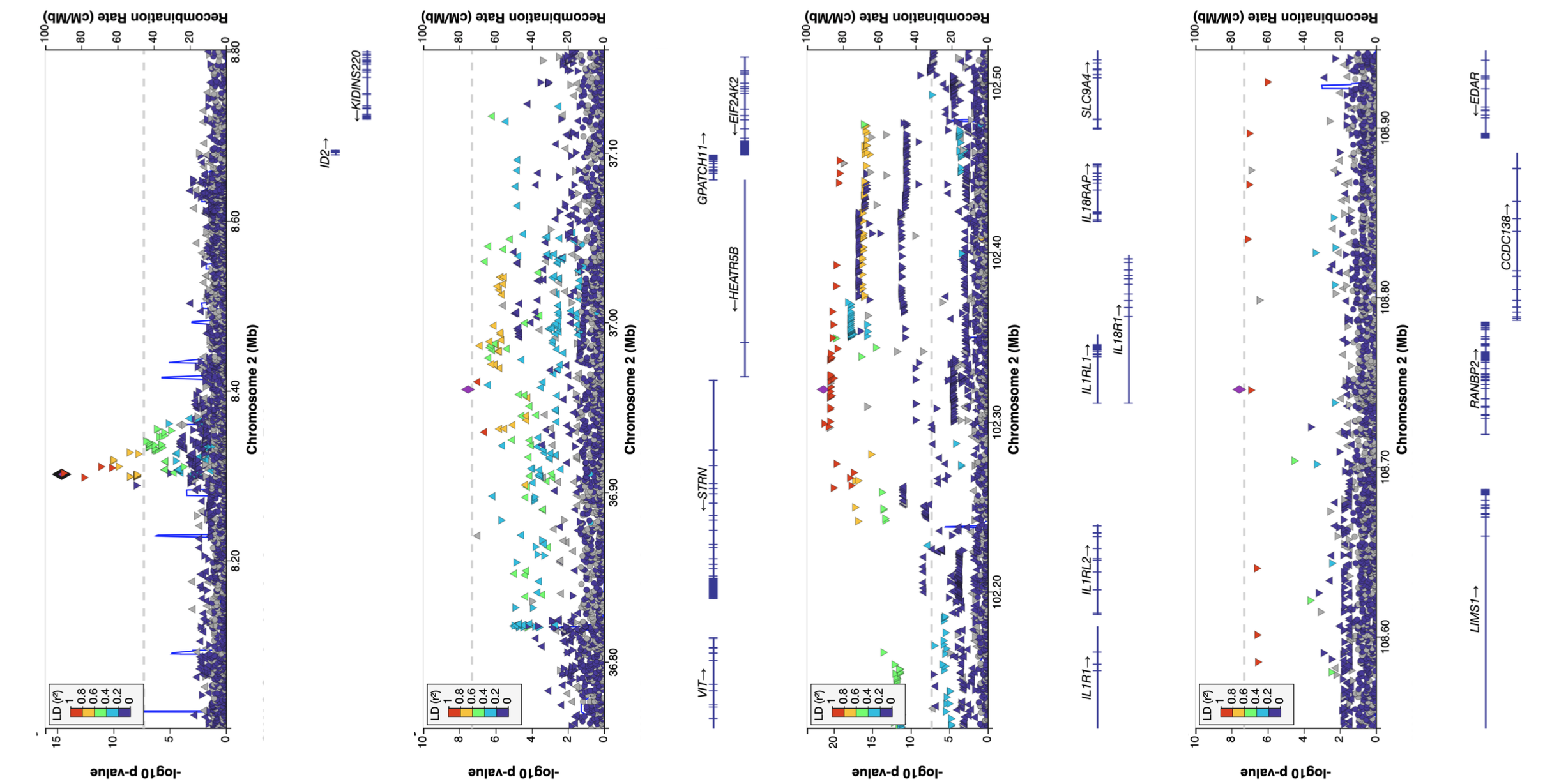

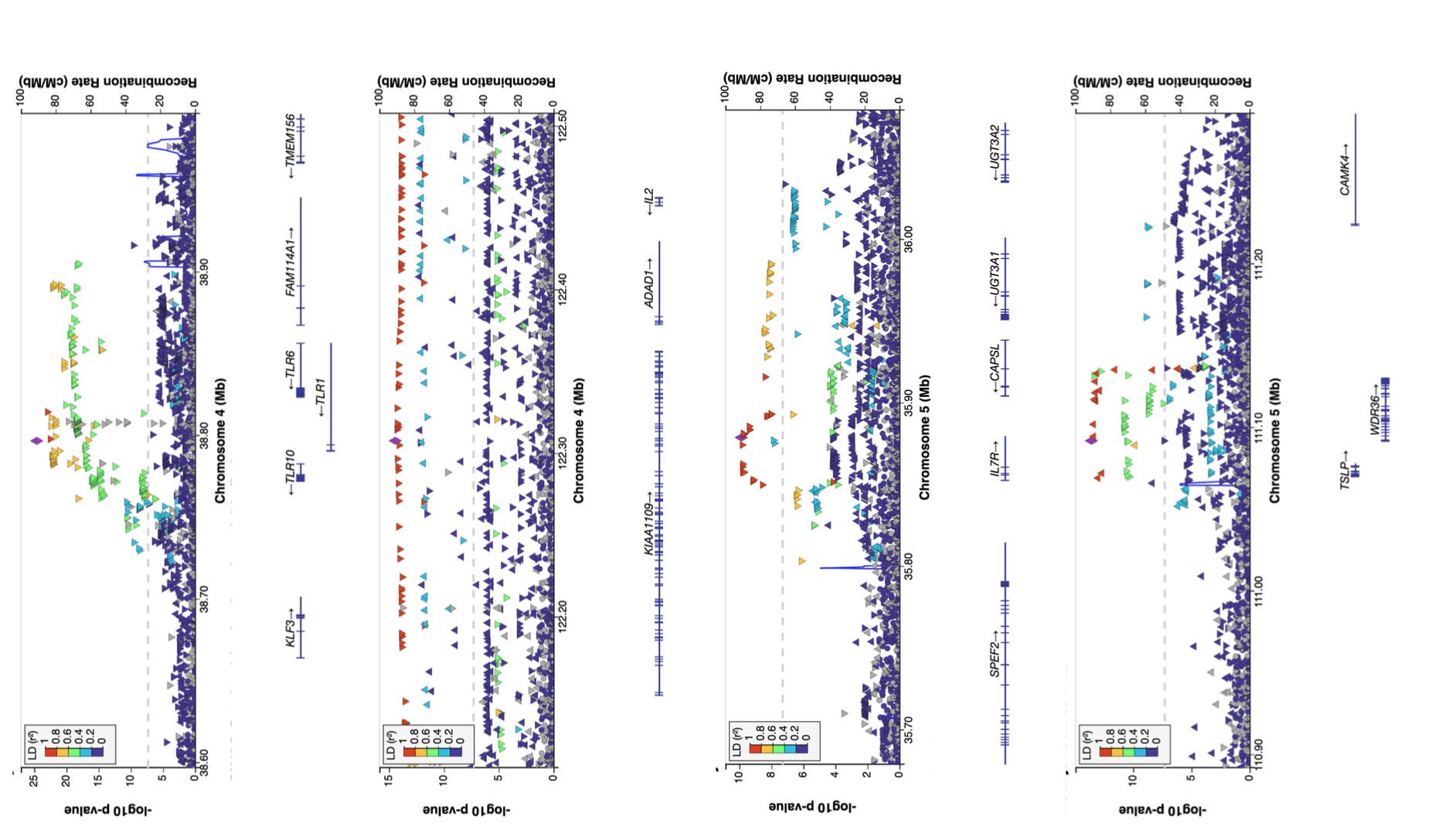

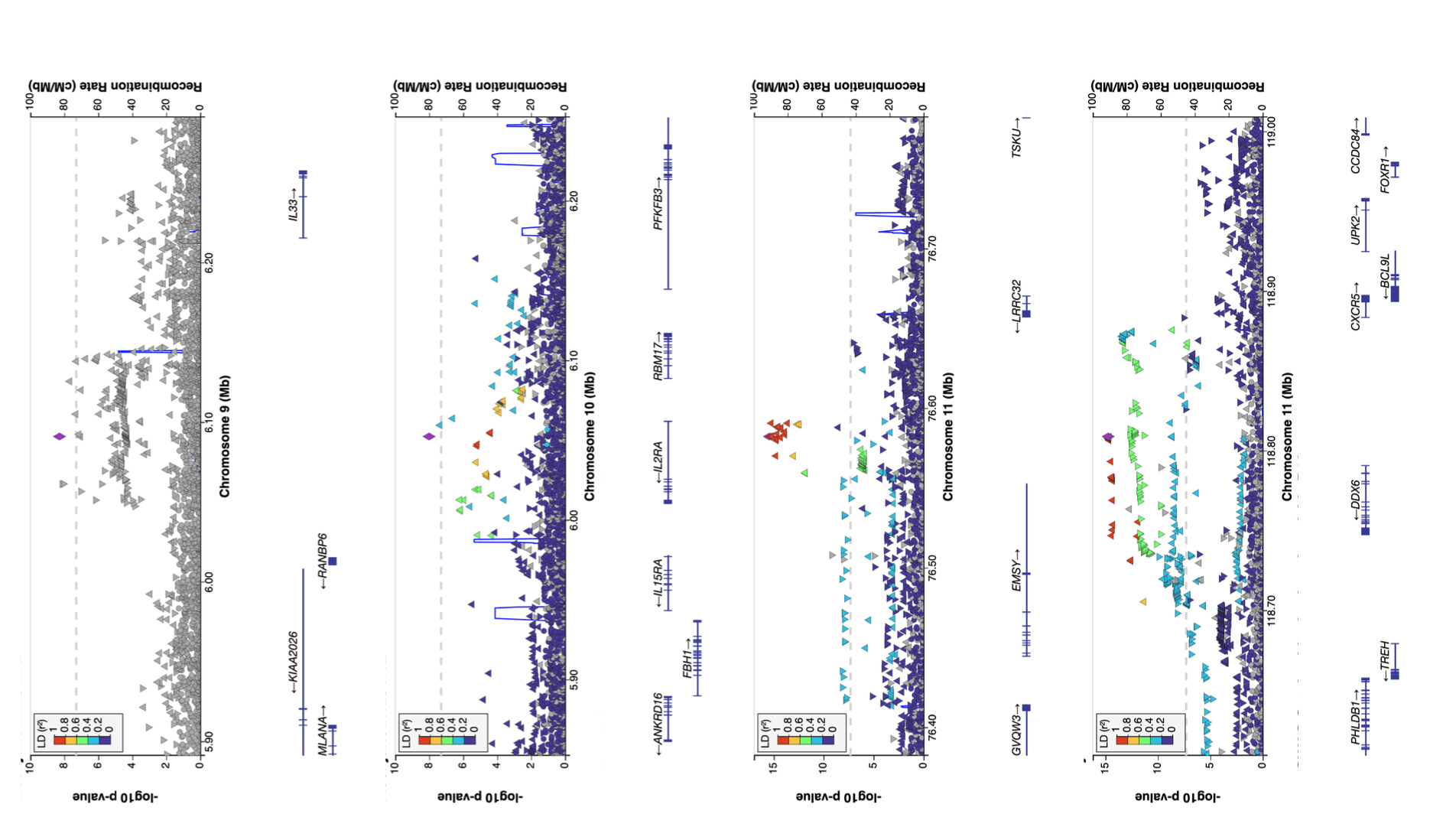

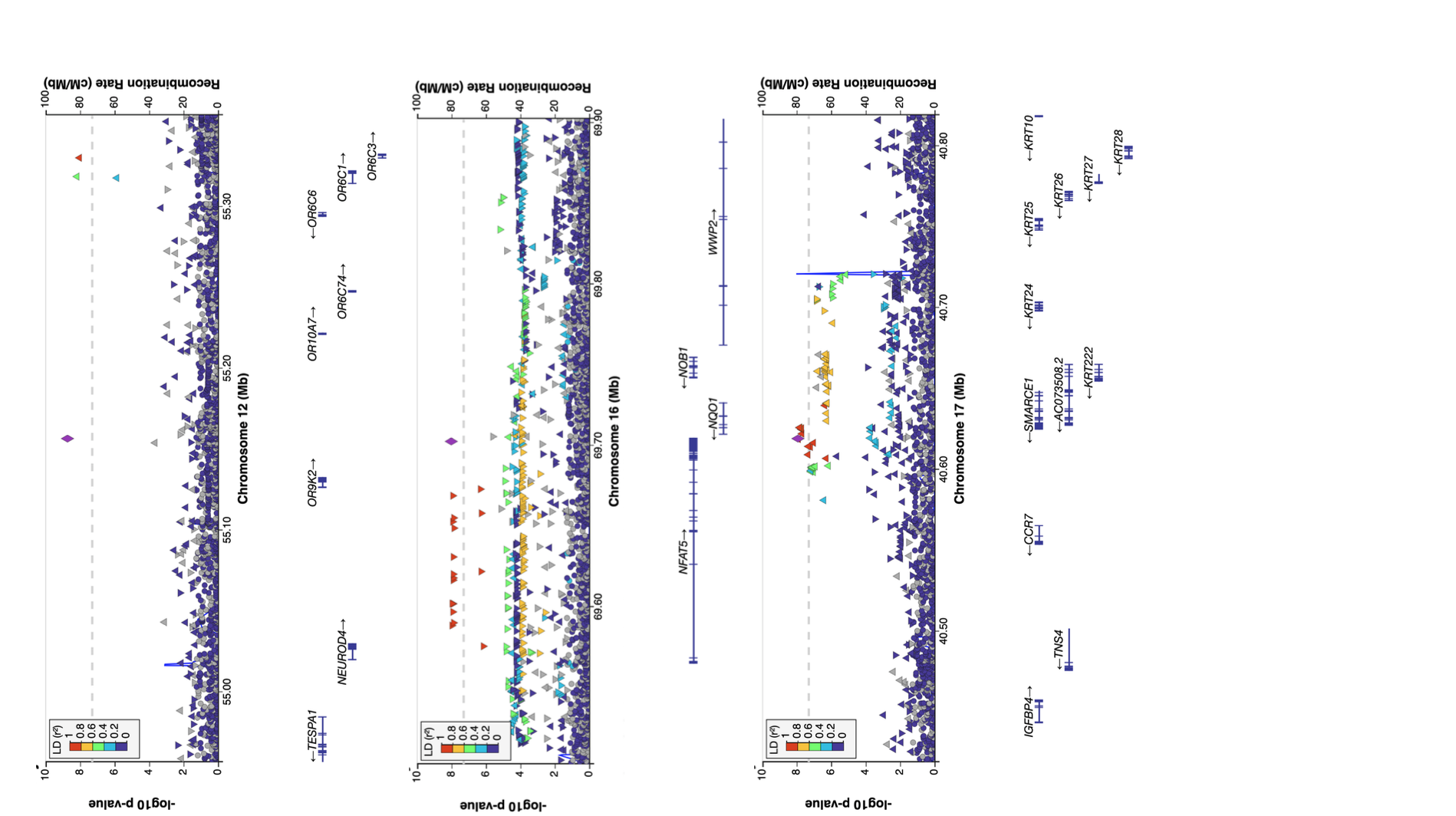

**Figure S2.** Regional association plots of selected genome-wide significant (p < 5 x 10-8) allergic conjunctivitis associated loci. European LD reference panel is applied. The genome-wide significance threshold is marked with a dashed grey line.

**SUPPLEMENTARY TEXT: ESTBB consortium authors**

Data collection, genotyping, QC and imputation:

Andres Metspalu -

Lili Milani -

Reedik Mägi -

Mait Metspalu

Mari Nelis -

Georgi Hudjashov -

Tõnu Esko -

**SUPPLEMENTAL MATERIAL REFERENCES**

1.	Kurki MI, Karjalainen J, Palta P, et al. FinnGen provides genetic insights from a well-phenotyped isolated population. *Nature*. 2023;613(7944):508-518. doi:10.1038/s41586-022-05473-8

2.	Leitsalu L, Haller T, Esko T, et al. Cohort Profile: Estonian Biobank of the Estonian Genome Center, University of Tartu. *Int J Epidemiol*. 2015;44(4):1137-1147. doi:10.1093/ije/dyt268

3.	Laisk T, Lepamets M, Koel M, Abner E, Estonian Biobank Research Team, Mägi R. Genome-wide association study identifies five risk loci for pernicious anemia. *Nat Commun*. 2021;12(1):3761. doi:10.1038/s41467-021-24051-6

4.	Sudlow C, Gallacher J, Allen N, et al. UK biobank: an open access resource for identifying the causes of a wide range of complex diseases of middle and old age. *PLoS Med*. 2015;12(3):e1001779. doi:10.1371/journal.pmed.1001779
